# Pre-stroke loop diuretics and anemia in elderly patients were associated with severe renal dysfunction at the onset of acute stroke

**DOI:** 10.1101/2023.07.10.23292484

**Authors:** Takahisa Mori, Tetsundo Yano, Kazuhiro Yoshioka, Yuichi Miyazaki

**Author notes:** **Corresponding author:** Takahisa Mori, MD, PhD, Department of Stroke Treatment, Shonan Kamakura General Hospital, Okamoto 1370-1, Kamakura City, Kanagawa 247-8533, Japan.

## Abstract

**Introduction:** Severe renal dysfunction (SRD) is a common complication of chronic kidney disease (CKD) that is a risk factor for heart disease and acute stroke (AS).

Furthermore, SRD can limit the treatment options for AS patients and influence their prognosis. Thus, preventing CKD progression to SRD and identifying the factors contributing to SRD at AS onset is crucial. However, the frequency of SRD in AS patients and the associated factors are poorly understood in different genders. In this study, we aimed to investigate the frequency of SRD in AS patients and analyze the associated factors by sex.

**Methods:** Our cross-sectional study included patients who met the following criteria: 1) admission within 24 hours of AS onset between 2013 and 2019 and 2) availability of pre-stroke medication information. We used the Cockcroft-Gault equation for calculating creatinine clearance (Ccr) and defined SRD as Ccr <30 ml/min. Then, we performed a multivariable logistic regression analysis to identify independent factors associated with SRD on admission separately for males and females.

**Results:** Out of 4294 patients with AS, 3472 were included for analysis. Of these, 1905 (54.9%) were male, with a median age of 75 and 81 years for males and females, respectively. The prevalence of SRD was 9.7% in males and 18.7% in females. For males, factors associated with SRD were loop diuretics, aspirin, L-type calcium channel blockers, alpha-beta blockers, and anemia. For females, factors associated with SRD were loop diuretics, mineralocorticoid receptor antagonists, and anemia.

**Conclusions:** Loop diuretic use and anemia before AS onset were associated with SRD in both males and females. To prevent SRD, individualized drug and anemia management are essential. Further prospective studies are warranted to confirm our findings and to elucidate the causal mechanism of SRD in patients with AS.

## Introduction

Chronic kidney disease (CKD) is associated with vascular risk factors ^1-3^, heart disease, and stroke ^4-11^. If CKD progresses to severe renal dysfunction (SRD) before the onset of acute stroke (AS), SRD can limit the treatment options for AS patients ^12-21^ and influence their prognosis. Therefore, preventing CKD progression to SRD is crucial for improving the outcomes of AS patients. Moreover, identifying the factors contributing to SRD at the onset of AS may help to optimize the management of AS patients.

However, the frequency of SRD in AS patients and the associated factors are poorly understood, especially by sex. In this study, we aimed to investigate the frequency of SRD in AS patients and analyze the associated factors by sex.

## Methods

### Patients

We conducted a cross-sectional study based on an institutional stroke registry database with a prospective and consecutive enrollment of patients. We included patients who met the following criteria: 1) admission within 24 hours of AS onset between January 2013 and March 2019 and 2) availability of pre-stroke medication information. We excluded patients whose serum creatinine level or body weight was not measured at arrival.

### Materials

We collected data at arrival on age; sex; body weight; pre-stroke medications; serum levels of albumin, glucose, glycated hemoglobin A1c, lipids, and c-reactive protein; pre-stroke modified Rankin scale; and stroke subtypes (ischemia or hemorrhage). Lipids were total cholesterol, low-density lipoprotein cholesterol, high-density lipoprotein cholesterol, and triglycerides. Antihypertensives were alpha (α) blockers, alpha-beta (αβ) blockers, beta (β) blockers, angiotensin-converting enzyme inhibitors (ACEis), angiotensin receptor blockers, L-type calcium channel blockers (CCBs), T-type or

N-type CCBs, and mineralocorticoid receptor antagonists (MRAs). Diabetes medications were α glucosidase inhibitors (αGIs), dipeptidyl peptidase 4 inhibitors, biguanides, sodium-glucose cotransporter 2 inhibitors, insulin secretagogues (ISs), and long-acting insulin analogs. Diuretics were loop or thiazide. Antiplatelets were aspirin or non-aspirins. Anticoagulants were warfarin or direct-acting oral anticoagulants (DOACs).

### Procedures

We used the Cockcroft-Gault equation for calculating creatinine clearance (Ccr) using age, body weight, serum creatinine level, and sex ^22^. Ccr is correlated with glomerular filtration rate ^23^, and Ccr <30 ml/min limits pharmacological treatments ^14,15,18-20^.

Therefore, we defined SRD as Ccr <30 ml/min and investigated the frequency of SRD in AS patients and significant factors for SRD by sex.

### Ethics approval and consent to participate

All procedures were performed in accordance with the ethical standards of the institution and the 1964 Declaration of Helsinki. The Tokushukai Group Ethics Committee approved our retrospective study (TGE01985-024). Furthermore, the Tokushukai Group Ethics Committee waived the need for written informed consent as the enrollment of study participants was based on an opt-out model.

### Statistical analysis

We expressed distributed continuous variables as median and interquartile ranges. We used a dummy variable (1 or 0) to represent categorical data, such as data on sex, for multivariable analysis. We performed the multivariable logistic regression analysis to identify independent factors associated with SRD at admission separately for males and females. We excluded age, body weight, and serum creatinine level from the multivariable analysis since they are used to calculate Ccr. A *p-value* <0.05 was considered statistically significant. We used the JMP software (version 17.1; SAS Institute, Cary, NC, USA) for all statistical analyses. One author (TM) had full access to all the data in the study and took responsibility for its integrity and data analysis.

## Results

Out of 4294 patients with AS, 3472 were included in the analysis. Of these, 1905 (54.9%) were male, with a median age of 75 years for males and 81 years for females. The prevalence of SRD was 9.7% in males and 18.7% in females (Table 1), which shows the baseline characteristic of the patients stratified by sex. The medications before stroke onset are summarized for males and females in supplemental Table 1.

**Table 1.** Characteristics for males and females on admission

| variables | Male sex | Female sex |
| --- | --- | --- |
| number | 1905 | 1567 |
| Ccr <30 ml/min on admission, n (%) | 185 (9.7%) | 293 (18.7%) |
| Age, years | 75, 66.5–82 | 81, 72–87 |
| Modified Rankin Scale before the acute stroke | 0, 0–2 | 0, 0–3 |
| Ischemia, n (%) | 1517 (79.6%) | 1218 (77.7%) |
| Body Mass Index, kg/m<sup>2</sup> | 22.9, 20.8–25.0 | 20.9, 18.7–23.6 |
| Height, cm | 165, 161–170 | 151.4, 148–156 |
| Body weight, kg | 62.9, 55.9–70 | 48.4, 41.5–55 |
| Albumin, g/L | 40, 37–43 | 40, 36–43 |
| Hemoglobin, g/L | 141, 128–152 | 129, 117–1139 |
| Glucose, mmol/L | 6.88, 5.83–8.66 | 6.61, 5.77–8.27 |
| Glycated hemoglobin A1c, % | 5.8, 5.5–6.4 | 5.8, 5.5–6.2 |
| Low-density lipoprotein cholesterol, mmol/L | 2.81, 2.22–3.40 | 3.19, 2.58–3.86 |
| High-density lipoprotein cholesterol, mmol/L | 1.35, 1.11–1.66 | 1.56, 1.29–1.90 |
| Triglyceride, mmol/L | 1.12, 0.78–1.76 | 1.04, 0.77–1.48 |
| Creatinine, µmol/L | 82.2, 70.7–100.3 | 61.9, 53.0–73.4 |
| C-reactive protein, µg/L | 1200, 500–3900 | 1200, 500–4700 |

All values except for categorical data are represented as median and interquartile ranges.
Ccr, creatinine clearance

For males, multivariable logistic regression analysis identified loop diuretics, aspirin, L-type CCBs, αβ blockers, α blockers, αGI, anemia, low albumin, and high glucose levels as independent factors that increased the risk of SRD, while the ACEis, ISs, and DOACs as factors that decreased the risk of SRD (Table 2). Moreover, the operating characteristics curve of logistic regression analysis revealed that the cut-off values of hemoglobin, albumin, and glucose for predicting Ccr <30 ml/min in male patients were ≤130 g/L (13.0 g/dL), ≤38 g/L (3.8 mg/dL), and ≥8.38 mmol/L (151.0 mg/dL), respectively (supplemental Tables 2-4). For females, multivariable logistic regression analysis identified loop diuretics, anemia, and MRAs as independent factors that increased the risk of SRD, while DOACs as factors that decreased the risk of SRD (Table 3). Moreover, the operating characteristics curve of logistic regression analysis revealed that the cut-off value of hemoglobin for predicting Ccr <30 ml/min in female patients was ≤124 g/L (supplemental Table 5).

**Table 2.** Multivariable logistic regression analysis for Ccr <30 ml/min in male patients

| Variables | OR | 95% CI | p-value | AUC |
| --- | --- | --- | --- | --- |
|  |  |  | <0.0001 | 0.890 |
| Hemoglobin, g/L | 0.94, 0.93–0.96 |  | <0.0001 |  |
| Loop diuretics (1:use, 0:no use) | 3.98, 2.38–6.63 |  | <0.0001 |  |
| ACEi (1:use, 0:no use) | 0.26, 0.10–0.62 |  | 0.0017 |  |
| Modified Rankin scale before the acute stroke | 1.21, 1.07–1.37 |  | 0.0025 |  |
| Aspirin (1:use, 0:no use) | 1.84, 1.19–2.81 |  | 0.0059 |  |
| alpha-beta blockers (1:use, 0:no use) | 2.08, 1.19–3.62 |  | 0.0109 |  |
| DOAC (1:use, 0:no use) | 0.31, 0.09–0.79 |  | 0.0131 |  |
| Albumin, g/L | 0.95, 0.91–0.99 |  | 0.0133 |  |
| L-type CCB (1:use, 0:no use) | 1.73, 1.12–2.66 |  | 0.0143 |  |
| Insulin secretagogues (1:use, 0:no use) | 0.33, 0.11–0.84 |  | 0.0189 |  |
| $\alpha$ glucosidase inhibitors (1:use, 0:no use) | 3.13, 1.19–7.95 | | 0.0216 | |
| Glucose, mmol/L | 1.09, 1.01–1.17 |  | 0.0258 |  |
| alpha-blockers (1:use, 0:no use) | 3.24, 1.12–8.85 |  | 0.0299 |  |
| Statin (1:use, 0:no use) | 0.62, 0.36–1.02 |  | 0.0622 |  |
| beta-blockers (1:use, 0:no use) | 1.89, 0.97–3.60 |  | 0.0624 |  |
| thiazide diuretics (1:use, 0:no use) | 2.06, 0.97–3.60 |  | >0.100 |  |
| biguanide (1:use, 0:no use) | 1.62, 0.68–3.77 |  | >0.100 |  |
| Low-density lipoprotein cholesterol, mmol/L | 1.12, 0.87–1.42 |  | >0.100 |  |
| C-reactive protein, μg/L | 1.00, 0.99–1.00 | | >0.100 | |
| T-type or N-type CCB (1:use, 0:no use) | 1.42, 0.53–3.46 |  | >0.100 |  |
| Glycated hemoglobin A1c, % | 0.93, 0.75–1.15 |  | >0.100 |  |
| diltiazem (1:use, 0:no use) | 0.53, 0.03–3.13 |  | >0.100 |  |
| MRA (1:use, 0:no use) | 0.73, 0.15–2.50 |  | >0.100 |  |
| High-density lipoprotein cholesterol, mmol/L | 0.89, 0.54–1.45 |  | >0.100 |  |
| Long-acting insulin analogs (1:use, 0:no use) | 0.82, 0.29–2.11 |  | >0.100 |  |
| warfarin (1:use, 0:no use) | 0.90, 0.46–1.68 |  | >0.100 |  |
| SGLT2i (1:use, 0:no use) | 0.73, 0.03–6.07 |  | >0.100 |  |
| Angiotensin receptor blockers (1:use, 0:no use) | 1.04, 0.67–1.60 |  | >0.100 |  |
| DPP4i (1:use, 0:no use) | 0.98, 0.49–1.91 |  | >0.100 |  |
| verapamil (1:use, 0:no use) | 0.97, 0.25–3.43 |  | >0.100 |  |
| Triglyceride, mmol/L | 0.99, 0.78–1.23 |  | >0.100 |  |
| Antiplatelets except aspirin (1:use, 0:no use) | 1.00, 0.58–1.70 |  | >0.100 |  |

ACEi, angiotensin-converting enzyme inhibitors; AUC, area under the curve; CCB, calcium channel blockers; CI, confidence interval; DOAC, direct-acting oral anticoagulants; DPP4i, dipeptidyl peptidase 4 inhibitors; MRA, mineralocorticoid receptor antagonists; OR, odds ratio; p, probability; SGLT2i, sodium-glucose co-transporter 2 inhibitors

**Table 3.** Multivariable logistic regression analysis for Ccr <30 ml/min in female patients

| Variables | OR | 95% CI | p-value | AUC |
| --- | --- | --- | --- | --- |
|  |  |  | <0.0001 | 0.815 |
| Loop diuretics (1:use, 0:no use) | 5.20, | 3.44–7.89 | <0.0001 |  |
| Hemoglobin, g/L | 0.96, | 0.95–0.97 | <0.0001 |  |
| Modified Rankin scale before the acute stroke | 1.26, | 1.15–1.39 | <0.0001 |  |
| DOAC (1:use, 0:no use) | 0.27, | 0.09–0.67 | 0.0031 |  |
| MRA (1:use, 0:no use) | 2.37, | 1.12–4.88 | 0.0249 |  |
| L-type CCB (1:use, 0:no use) | 1.43, | 0.99–2.05 | 0.0580 |  |
| alpha-beta blockers (1:use, 0:no use) | 1.62, | 0.95–2.74 | 0.0779 |  |
| thiazide diuretics (1:use, 0:no use) | 2.39, | 0.88–5.89 | 0.0847 |  |
| Insulin secretagogues (1:use, 0:no use) | 1.99, | 0.82–4.69 | >0.100 |  |
| Long-acting insulin analogs (1:use, 0:no use) | 2.58, | 0.74–7.93 | >0.100 |  |
| Low-density lipoprotein cholesterol, mmol/L | 0.89, | 0.74–1.05 | >0.100 |  |
| Verapamil (1:use, 0:no use) | 0.50, | 0.16–1.32 | >0.100 |  |
| Glycated hemoglobin A1c, % | 0.90, | 0.78–1.05 | >0.100 |  |
| High-density lipoprotein cholesterol, mmol/L | 1.28, | 0.89–1.83 | >0.100 |  |
| DPP4i (1:use, 0:no use) | 0.62, | 0.29–1.27 | >0.100 |  |
| Angiotensin receptor blockers (1:use, 0:no use) | 1.27, | 0.88–1.82 | >0.100 |  |
| Albumin, g/L | 0.98, | 0.94–1.01 | >0.100 |  |
| Antiplatelets except aspirin (1:use, 0:no use) | 0.72, | 0.42–1.20 | >0.100 |  |
| beta blockers (1:use, 0:no use) | 1.37, | 0.78–2.36 | >0.100 |  |
| Glucose, mmol/L | 0.96, | 0.89–1.03 | >0.100 |  |
| ACEi (1:use, 0:no use) | 1.41, | 0.75–2.55 | >0.100 |  |
| $\alpha$ glucosidase inhibitors (1:use, 0:no use) | 0.54 | 0.11–1.95 | >0.100 | |
| Aspirin (1:use, 0:no use) | 1.21, | 0.80–1.81 | >0.100 |  |
| SGLT2i (1:use, 0:no use) | 2.54e-9, | 0 | >0.100 |  |
| warfarin (1:use, 0:no use) | 1.13, | 0.65–1.93 | >0.100 |  |
| alpha-blockers (1:use, 0:no use) | 1.23, | 0.40–3.43 | >0.100 |  |
| Triglyceride, mmol/L | 1.03, | 0.83–1.25 | >0.100 |  |
| T-type or N-type CCB (1:use, 0:no use) | 1.10, | 0.53–2.14 | >0.100 |  |
| diltiazem (1:use, 0:no use) | 0.88, | 0.17–3.61 | >0.100 |  |
| biguanide (1:use, 0:no use) | 0.96, | 0.34–2.57 | >0.100 |  |
| C-reactive protein, μg/L | 1.00, | 0.99–1.00 | >0.100 | |
| Statin (1:use, 0:no use) | 1.01, | 0.68–1.49 | >0.100 |  |

ACEi, angiotensin-converting enzyme inhibitors; AUC, area under the curve; CCB, calcium channel blockers; CI, confidence interval; DOAC, direct-acting oral anticoagulants; DPP4i, dipeptidyl peptidase 4 inhibitors; MRA, mineralocorticoid receptor antagonists; OR, odds ratio; p, probability; SGLT2i, sodium-glucose co-transporter 2 inhibitors

## Discussion

Our findings indicate that the use of loop diuretics and the presence of anemia before the onset of AS increase the risk of SRD in both males and females. On the other hand, DOACs decrease the risk. Loop diuretics are commonly prescribed medications for congestive heart failure or hypertension, suggesting that preventing the progression of cardio-renal anemia (CRA) syndrome ^24^ is crucial.

The CRA syndrome suggests an interaction between anemia, congestive heart failure, and chronic kidney insufficiency, contributing to the deterioration of the anemia and cardiac and renal function ^24^. Even mild anemia, indicated by Hb <13.8 g/dL, increases the risk for progression to end-stage renal disease ^25^. Additionally, subjects with low hematocrits, <40% for men and <35% for women, have a significantly increased risk of end-stage renal disease^26^. Therefore, it is essential to treat anemia before the CRA syndrome progresses.

Early initiation of erythropoietin in pre-dialysis patients with non-severe anemia significantly slows the progression of renal disease and delays the initiation of renal replacement therapy ^27^. Moreover, treating anemia with erythrolein-stimulating agents improves energy and physical function in non-dialysis CKD patients ^28^. However, the optimal timing of initiating anemia treatment remains controversial.

Maintaining higher Hb levels (11.0 ≤ Hb < 13.0 g/dL) with darbepoetin alfa has been shown to better preserve renal function in patients with CKD not on dialysis compared to maintaining lower Hb levels (9.0≤ Hb <11.0 g/dL) ^29^. Therefore, maintaining high Hb levels is recommended before the progression of anemia. However, a high target hemoglobin level may not always benefit patients with CRA syndrome. A target hemoglobin level of 13.5 g/dL, compared with 11.3 g/dL, was associated with an increased risk and no incremental improvement in the quality of life ^30^. Therefore, in real-world clinical practice, initiating treatment of patients with Hb >130 g/L and renal function of Ccr ≥30 ml/min may be delayed and challenging, as our study found that Hb cut-off values of 130 g/L and 124 g/L were associated with predicting Ccr <30 ml/min in our male and female patients, respectively. This delay in treatment initiation can potentially lead to the progression of CRA syndrome by exacerbating the anemia and cardiac and renal function.

Furthermore, heart failure is also a risk of all stroke subtypes ^31^. Loop diuretics, commonly employed medications for congestive heart failure, have been reported to exacerbate renal function ^32^. This adverse effect is consistent with our finding that loop diuretic use was associated with SRD. In addition, αβ blockers, or aspirin, used in male patients in our study, are commonly prescribed medications for congestive heart failure, hypertension, or ischemic heart disease. Moreover, L-type CCBs like amlodipine are widely used for hypertension, a significant risk factor for CKD. However, in renal protective effects, L-type CCB is inferior to N-type or L-type CCB ^33,34^, suggesting consistency with our finding that L-type CCB was associated with SRD in our male patients.

On the other hand, DOACs have been reported to have anti-inflammatory activity and vascular protection ^35,36^, suggesting consistency with our finding that DOACs were inversely associated with SRD in both males and females.

Therefore, our study highlights the importance of considering CRA syndrome in patients with multiple vascular risk factors and suggests the need for individualized drug and anemia management to prevent SRD and improve the outcomes of AS patients.

### Limitations

Our study has several limitations. Firstly, the cross-sectional design may introduce reverse causality in the relationships of SRD with pre-stroke medications, anemia, and DOACs. Second, the study population was mainly elderly Japanese, which limits the generalizability of the study outcomes to non-Japanese populations due to potential racial differences in medication efficacy. Third, the history of drugs may have been misclassified, introducing information bias. Fourth, there might be some unknown confounders for SRD. Fifth, we did not collect data on basic demographic information (education, income, marriage), lifestyles (smoking, drinking, physical activity, energy intake, salt intake, carbohydrates intake, fat intake, percentages of saturated fatty acids, n-6 polyunsaturated fatty acids, and n-3 polyunsaturated fatty acids), and family history of stroke, which could have biased the results. Therefore, a prospective study is warranted to confirm our findings.

## Conclusion

Loop diuretics and anemia before the onset of AS were associated with SRD in males and females, whereas DOACs decreased the risk. In males, several medications were associated with SRD. To prevent SRD, individualized drug and anemia management are essential. Further prospective studies are warranted to confirm our findings and to establish how to prevent SRD in patients with AS.

## Data Availability

The datasets analyzed during the current study are available from the corresponding author upon reasonable request.

## Acknowledgments

We thank our co-medical staff for specialized assistance at our comprehensive stroke center.

